# Correlates of Adherence to the 2017 Clinical Practice Guidelines for Pediatric Hypertension in Safety-Net Clinics: A Two-Year Cross-Sectional Study

**DOI:** 10.1101/2022.10.03.22280528

**Authors:** Allison J. Carroll, Yacob G. Tedla, Roxane Padilla, Arjit Jain, Eduardo Segovia, Anoosh Moin, Andrea S. Wallace, Olutobi A. Sanuade, Craig B. Langman, Nivedita Mohanty, Justin D. Smith

## Abstract

**Importance:** The 2017 Clinical Practice Guideline (CPG) has categorized a greater proportion of children with elevated blood pressure (BP) or pHTN, and yet several barriers to CPG adherence have been noted.

**Objective:** To assess adherence to the 2017 CPG for the diagnosis and management of pHTN.

**Design:** Cross-sectional study using electronic health record-extracted data (January 1, 2018 to December 31, 2020).

**Setting:** AllianceChicago, a national Health Center Controlled Network of federally qualified health centers.

**Participants:** Children and adolescents (ages ≥3 and <18) who attended ≥1 visit and had ≥1 BP reading ≥90^th^ percentile or diagnosis of elevated BP or pHTN.

**Exposure:** BP ≥90^th^ percentile or ≥95^th^ percentile.

**Main Outcomes and Measures:** 1) diagnosis of pHTN or elevated BP, 2) BP management (antihypertensive medication, lifestyle counseling, referral), and 3) follow-up visit attendance. Descriptive statistics described the sample and rates of guideline adherence. Logistic regression analyses identified patient-and clinic-level predictors of primary outcomes.

**Results:** Guideline-adherent diagnosis was observed in 8,811/23,334 (37.8%) children with BP ≥90^th^ percentile, 6,427/15,423 (41.6%) children with BP ≥95^th^ percentile, and 146/2,542 (5.7%) children with ≥3 visits with BP ≥95^th^ percentile. A clinical decision support tool was used to calculate BP percentiles in 45.1% of cases and was associated with significantly greater odds of pHTN diagnosis (OR: 6.18, 95%CI: 5.06, 9.40). Among children with BP ≥95^th^ percentile, antihypertensive medication was prescribed to 795/15,422 (5.2%) children, lifestyle counseling was provided to 14,841/15,422 (96.2%), and a BP-related referral was given to 848/15,422 (5.5%). Children seen at clinics in rural versus urban settings were more likely to be prescribed antihypertensive medication (OR: 1.96, 95%CI: 1.59, 2.41) and less likely to be given a BP-related referral (OR: 0.01, 95%CI: 0.00, 0.06). Guideline-adherent follow-up was observed in 8,651/19,049 (45.4%) children with BP ≥90^th^ percentile and 2,598/15,164 (17.1%) children with BP ≥95^th^ percentile.

**Conclusions and Relevance:** Fewer than 50% of children with elevated BP had a guideline-adherent diagnosis code or attended guideline-adherent follow-up. Using the clinical decision support tool increased guideline-adherent diagnosis, but was underutilized. Further work is needed to understand how to best support implementation of tools promoting pHTN diagnosis, management, and follow-up.

**Key points:** *Question:* To what extent are clinicians adhering to the 2017 Clinical Practice Guideline for pediatric hypertension diagnosis, management, and follow-up?

*Findings:* In this cross-sectional study of 23,334 children (3-17 years old) with elevated blood pressure, less than half of children had a corresponding diagnosis in their medical chart or attended the recommended follow-up visit. A clinical decision support tool that aided in classifying blood pressure values increased rates of diagnosis, but was underutilized.

*Meaning:* Findings suggest that pediatric hypertension and elevated blood pressure remain underdiagnosed and undertreated among high-risk children, which portends negative health consequences in adulthood.

## Background

The prevalence of pediatric hypertension (pHTN) is estimated to be approximately 4% globally.^1^ The prevalence rate of pHTN has risen in recent years,^2^ likely due to multiple factors (e.g., high rates of overweight and obesity).^2,3^ Importantly, this rise is due in part to the American Academy of Pediatrics 2017 Clinical Practice Guideline (CPG) for the diagnosis and management of pHTN,^4,5^ which replaced the 2004 CPG (i.e., the “Fourth Report”).^6^ According to one analysis of children (8-17 years old), 1.5 to 2.5% more children met criteria for pHTN per the 2017 CPG versus the Fourth Report.^7^ Compared to the Fourth Report, diagnosis adhering to the 2017 CPG significantly improved prediction of adult HTN from 13% to 22%.^8^

Prior investigations have demonstrated that children with elevated BP/pHTN frequently go undiagnosed.^9-11^ For example, one study found that only 14.6% of children who met diagnostic criteria (per the Fourth Report) for elevated blood pressure (BP) or pHTN per elevated BP measurements had a corresponding ICD-10 diagnosis code in the electronic health record (EHR).^12^ Another EHR study found that although BP was nearly always measured at well-child visits, less than half of children with elevated BP received appropriate, guideline-adherent management.^10^ Accurate diagnosis of elevated BP and pHTN is critical for obtaining appropriate treatment to reduce the risk of future health consequences. For example, elevated BP in children is associated with impairments in cardiac (e.g., increased left ventricular mass) and vascular (e.g., increased arterial stiffness) structures^8,13,14^ as well as neurocognitive and psychological effects.^15^ Consequently, children with pHTN are at greater risk of developing hypertension, metabolic syndrome, and left ventricular hypertrophy in adulthood.^16,17^

Numerous barriers to appropriate pHTN diagnosis and management have been identified, including high patient volumes and short visit times in primary care; low confidence in BP readings due to poor measurement technique or inadequate equipment; and lack of appropriate follow-up by patients.^18,19^ A common strategy employed to try to overcome these barriers is the use of clinical decision support (CDS) tools in the EHR to save time and decrease clinician decision-making burden, and have been shown to improve recognition of pHTN.^20,21^

To date, most studies examining pHTN guideline adherence, as well as use of CDS tools, have been conducted using the guidelines in the Fourth Report. The purpose of this study was to examine the rates of adherence to the 2017 CPG, which also included implementation of a CDS tool in the EHR designed to facilitate guideline adherence. Specifically, we examined the rates and predictors of guideline-adherent pHTN diagnosis, management (antihypertensive medication, lifestyle counseling, referral), and follow-up visits. We also examined whether clinicians were using the CDS and if use of the CDS increased the likelihood of achieving guideline-adherent pHTN diagnosis, management, and follow-up visits.

## Methods

### Setting

This study extracted data from the EHR of AllianceChicago, a Health Center Controlled Network of federally qualified health centers (FQHCs; N=72); the dataset in this study represents FQHCs in 17 states. The FQHCs in the network provide services for communities in urban, suburban, and rural settings. Patients seen at FQHCs within the AllianceChicago network come from primarily uninsured, underinsured, and low-income populations. The FQHCs are located in areas that have high African American and Hispanic/Latino populations.

### Study period

Data from visits that occurred between January 1, 2018 and December 31, 2019 were retrieved for this study. This time frame was chosen because the recommendations from the 2017 CPG for HTN were implemented through health information technology tools throughout the AllianceChicago network, including an EHR-based CDS to automatically calculate BP percentiles and classify blood pressure values, starting January 1, 2018.

### Participants

This study included 23,334 children and adolescents (ages ≥3 and <18) who attended at least one healthcare visit in the AllianceChicago network during the study period, and had 1) ≥1 systolic or diastolic BP reading ≥90^th^ percentile during the study period and/or 2) ICD-10 diagnosis for elevated BP (R03.0: Elevated blood-pressure reading, without diagnosis of hypertension) or pHTN (I10: Essential (Primary) Hypertension). Patients were excluded if they were pregnant or diagnosed with heart disease.

### Data collection

Participants in the AllianceChicago network use a centrally hosted, common EHR system (AthenaPractice). All visit information, patient characteristics, vital signs, and orders, are recorded in the EHR.

#### Patient and clinic characteristics

Patient characteristics included sex (male or female), age (3 to 6 years, 7 to 11 years, or 12 to 17 years), race (Asian, Black/African American, Hispanic/Latino, White, Other, or Unknown), ethnicity (Hispanic/Latino, Not Hispanic/Latino, or Unknown), and weight status per body mass index percentile (<5% [Underweight], ≥5 and <85% [Normal Weight], ≥85 and <95% [Overweight], or ≥95% [Obese]). The clinic-level characteristic included the setting of the clinic (Urban, Suburban, Rural, or Unknown).

#### CDS tool use

The CDS was designed to automate calculation of BP percentiles, classify BP values and identify abnormal values, provide guidance on obtaining manual BP values, and calculate averages per the 2017 CPG. The CDS tool was coded 1 if the clinician used the tool to assign a BP percentile at all visits with an available BP value, and coded as 0 if the clinician did not use the tool at ≥1 visits with an available BP value.

#### Guideline-adherent diagnosis

A guideline-adherent diagnosis was coded if:

1. Child had ≥1 systolic or diastolic BP ≥90^th^ percentile if aged <13, or systolic BP ≥120 mmHg or diastolic BP ≥80 mmHg if aged ≥13, and an elevated BP diagnosis (R03.0)
2. Child had ≥1 systolic or diastolic BP ≥95^th^ percentile if aged <13, or systolic BP ≥130 mmHg or diastolic BP ≥80 mmHg if aged ≥13, and an elevated BP diagnosis (R03.0)
3. Child had ≥3 systolic or diastolic BP ≥95^th^ percentile if aged <13, or systolic BP ≥130 mmHg or diastolic BP ≥80 mmHg if aged ≥13, and a pediatric hypertension diagnosis (I10).

#### Guideline-adherent management

The possible management options for elevated BP included 1) antihypertensive medications, 2) lifestyle counseling, and 3) referrals. First, we extracted data about whether or not a child was prescribed an antihypertensive medication (e.g., ACE inhibitors, beta blockers; see Supplemental Material for a complete list) at the time of the visit. Second, we extracted data about whether or not the child received lifestyle counseling (Z71.3: Dietary counseling and surveillance; Z71.82: Exercise counseling; or CPT codes 99401, 99402, 99403, 99404: Preventative medicine counseling and/or risk factor reduction intervention(s) provided to an individual) at the visit. Third, we extracted data about whether or not the child received a hypertension-relevant referral at the visit (e.g., cardiology, nephrology).

#### Guideline-adherent follow-up visits

The 2017 CPG recommends that patients who have a BP reading ≥90^th^ percentile attend a follow-up visit within 6 months and those with a BP reading ≥95^th^ percentile attend a follow-up visit within 2 weeks. Thus, patients with BP readings ≥90^th^ and <95^th^ percentile were eligible to be included in this analysis if they attended a first visit between January 1, 2018 and June 30, 2019, and were coded as attending guideline-adherent follow-up if they had a second visit within 6 months of the first. Patients with BP readings ≥95^th^ percentile were eligible to be included in this analysis if they attended a first visit between January 1, 2018 and Nov 30, 2019, and coded as attending guideline-adherent follow-up if they had a second visit within 1 month of the first (to conservatively allow for 2-week follow-up, consistent with prior studies^22^).

### Statistical analyses

Numbers and percentages were calculated to describe patients’ baseline characteristics for those who 1) had ≥1 BP ≥90^th^ percentile (full sample), 2) had ≥1 BP ≥95^th^ percentile, and 3) attended ≥3 visits. Second, we conducted multivariate logistic regressions for guideline-adherence outcomes. Guideline-adherent diagnosis models included 1) ≥1 BP ≥90^th^ percentile with elevated BP diagnosis, 2) ≥1 BP ≥95^th^ percentile with elevated BP diagnosis, and 3) ≥3 BP ≥95^th^ percentile with pHTN diagnosis. We also examined whether CDS tool use was associated with guideline-adherent diagnosis. Guideline-adherent management models included models for BP ≥90^th^ and BP ≥95^th^ percentile, each with 1) antihypertensive medications, 2) lifestyle counseling, and 3) referrals. Guideline-adherent follow-up models included 1) BP ≥90^th^ percentile with 6-month follow-up attendance and 2) BP ≥95^th^ percentile with 1-month follow-up attendance. Covariates in all models included patient sex (male or female), age (years), race (Asian, Black, Hispanic/Latino, White, More than One Race, Other, Unknown), ethnicity (Hispanic/Latino, Not Hispanic/Latino, Unknown), weight status (underweight, normal weight, overweight, obese), and clinic setting (urban, suburban, rural, unknown). All analyses were conducted using Stata 17.^23^ A 2-tailed *p*< .05 was considered statistically significant.

## Results

### Sample description

The full sample comprised 23,334 children with BP ≥90^th^ percentile on ≥1 visits. Of the full sample, 15,422 children (66%) had BP ≥95^th^ percentile on ≥1 visits and 11,675 children (50%) had BP measured on ≥3 visits. Characteristics of each sample are provided in Table 1.

**Table 1.** Sample characteristics of children (3-18 years old) with elevated blood pressure readings (January 1st, 2018 through December 31st, 2019)

| Variable | ≥1 elevated BP reading ≥90th percentile (N=23,334) |  | ≥1 elevated BP reading ≥95th percentile (n=15,422) |  | ≥1 elevated BP reading ≥90th percentile & attended ≥3 visits (n=11,675) |  |
| --- | --- | --- | --- | --- | --- | --- |
|  | N | % | n | % | n | % |
| Sex |  |  |  |  |  |  |
| Male | 12811 | 54.9% | 8563 | 55.5% | 6520 | 55.8% |
| Female | 10523 | 45.1% | 6859 | 44.5% | 5155 | 44.2% |
| Age |  |  |  |  |  |  |
| ≤6 years | 8139 | 34.9% | 5271 | 34.2% | 3343 | 28.6% |
| 7-11 years | 9356 | 40.1% | 5999 | 38.9% | 4931 | 42.2% |
| ≥12 years | 5839 | 25.0% | 4151 | 26.9% | 3401 | 29.1% |
| Race |  |  |  |  |  |  |
| Asian | 715 | 3.1% | 489 | 3.2% | 346 | 3.0% |
| Black | 6084 | 26.1% | 3970 | 25.7% | 2974 | 25.5% |
| Hispanic/Latino <sup>a</sup> | 1417 | 6.1% | 924 | 6.0% | 620 | 5.3% |
| White | 13663 | 58.6% | 9039 | 58.6% | 7159 | 61.3% |
| More than one race | 316 | 1.4% | 218 | 1.4% | 148 | 1.3% |
| Other | 284 | 1.2% | 197 | 1.3% | 116 | 1.0% |
| Unknown | 855 | 3.7% | 587 | 3.8% | 312 | 2.7% |
| Ethnicity |  |  |  |  |  |  |
| Hispanic/Latino | 11978 | 51.3% | 7836 | 50.8% | 6197 | 53.1% |
| Not Hispanic/Latino | 10687 | 45.8% | 7176 | 46.5% | 5205 | 44.6% |
| Unknown | 669 | 2.9% | 410 | 2.7% | 273 | 2.3% |
| Weight status |  |  |  |  |  |  |
| Underweight | 486 | 2.1% | 316 | 2.0% | 229 | 2.0% |
| Normal weight | 9727 | 41.7% | 6010 | 39.0% | 4518 | 38.7% |
| Overweight | 3800 | 16.3% | 2427 | 15.7% | 1888 | 16.2% |
| Obese | 9321 | 39.9% | 6669 | 43.2% | 5040 | 43.2% |
| Clinic setting |  |  |  |  |  |  |
| Urban | 16860 | 72.3% | 10915 | 70.8% | 8585 | 73.5% |
| Suburban | 3343 | 14.3% | 2275 | 14.8% | 1556 | 13.3% |
| Rural | 2673 | 11.5% | 1871 | 12.1% | 1324 | 11.3% |
| Unknown | 460 | 2.0% | 361 | 2.3% | 210 | 1.8% |

<sup>a</sup> The race category of "Hispanic/Latino" was included when it was entered as the Race in the electronic health record.

### Guideline-adherent diagnosis

Logistic regression analyses predicting guideline-adherent diagnosis are presented in Table 2.

**Table 2.** Predictors having a guideline-adherent ICD-10 diagnosis code for elevated blood pressure or pediatric hypertension in the electronic health record among children with elevated blood pressure readings

| Variable | BP ≥90th percentile at ≥1 visit<br>(N=23,334)<sup>a</sup> |  |  | BP ≥95th percentile at ≥1 visit<br>(N=15,423)<sup>b</sup> |  |  | BP ≥95th percentile at ≥3 visits<br>(N=2,542)<sup>c</sup> |  |  |
| --- | --- | --- | --- | --- | --- | --- | --- | --- | --- |
|  | OR | 95% CI |  | OR | 95% CI |  | OR | 95% CI |  |
| age, years | **1.264** | **1.255** | **1.274** | **1.259** | **1.247** | **1.270** | **1.024** | **1.018** | **1.031** |
| sex (ref: male) | 0.988 | 0.929 | 1.051 | 1.025 | 0.950 | 1.105 | 0.953 | 0.902 | 1.007 |
| Race (ref: non-Hispanic white) |  |  |  |  |  |  |  |  |  |
| Asian | 1.207 | 0.985 | 1.480 | **1.279** | **1.000** | **1.635** | 1.020 | 0.855 | 1.218 |
| Black | **1.240** | **1.117** | **1.375** | **1.302** | **1.149** | **1.477** | 0.970 | 0.886 | 1.063 |
| Hispanic/Latino | **0.469** | **0.409** | **0.538** | **0.458** | **0.388** | **0.540** | 0.975 | 0.867 | 1.097 |
| More than one race | 0.910 | 0.661 | 1.254 | 0.918 | 0.625 | 1.348 | 0.965 | 0.749 | 1.243 |
| Other | 0.731 | 0.506 | 1.056 | 0.914 | 0.595 | 1.404 | 0.795 | 0.600 | 1.055 |
| Unknown | **0.781** | **0.642** | **0.950** | **0.758** | **0.602** | **0.954** | **1.236** | **1.043** | **1.464** |
| Ethnicity (ref: not Hispanic/Latino) |  |  |  |  |  |  |  |  |  |
| Hispanic/Latino | **1.646** | **1.494** | **1.814** | **1.669** | **1.484** | **1.876** | 0.932 | 0.856 | 1.014 |
| Unknown | **1.220** | **1.001** | **1.486** | 1.194 | 0.933 | 1.528 | **0.718** | **0.607** | **0.851** |
| Weight status (normal BMI) |  |  |  |  |  |  |  |  |  |
| Underweight | 0.825 | 0.646 | 1.053 | 0.767 | 0.572 | 1.029 | 1.167 | 0.963 | 1.413 |
| Overweight | 1.078 | 0.985 | 1.180 | 1.049 | 0.938 | 1.174 | 1.080 | 0.998 | 1.168 |
| Obese | **1.242** | **1.158** | **1.331** | **1.267** | **1.164** | **1.380** | **1.488** | **1.397** | **1.585** |
| Clinic setting (ref: Urban) |  |  |  |  |  |  |  |  |  |
| Suburban | **0.589** | **0.539** | **0.645** | **0.603** | **0.542** | **0.671** | **1.167** | **1.077** | **1.265** |
| Rural | **0.254** | **0.224** | **0.289** | **0.243** | **0.209** | **0.282** | **1.266** | **1.146** | **1.399** |
| Unknown | **0.291** | **0.212** | **0.401** | **0.274** | **0.192** | **0.391** | **2.325** | **1.799** | **3.005** |
| Constant | **0.067** | **0.058** | **0.078** | **0.078** | **0.064** | **0.093** | **1.430** | **1.258** | **1.627** |

**Bold** indicates a significant predictor in a logistic regression predicting that eligible children had an ICD-10 diagnosis code.
<sup>a</sup> 8,810 out of 23,334 (37.8%) eligible children had a diagnosis of elevated BP in the EHR.
<sup>b</sup> 6,425 out of 15,423 (41.7%) eligible children had diagnosis of elevated BP in the EHR.
<sup>c</sup> 146 out of 2,542 (5.7%) eligible children had a diagnosis of pHTN in the EHR.
Abbreviations: BP: blood pressure. EHR: electronic health record. pHTN: pediatric hypertension. OR: odds ratio. CI: confidence interval. BMI: body mass index.

All children in the present sample (N=23,334) met criteria for elevated BP (i.e., had a BP reading ≥90^th^ percentile at ≥1 visits), but only 8,811 (37.8%) had a corresponding ICD-10 code (R03.0). Those children with BP ≥90^th^ percentile who had the diagnosis code, versus those who did not, were more likely to be older (77.7% of children ≥12 years vs. 23.3% of children ≤6 years), Black (40.7%) versus white race (38.9%) and white versus Hispanic/Latino race (31.2%), Hispanic/Latino (42.2%) or unknown (35.7%) versus white ethnicity (32.9%), classified with obesity (46.5%) versus normal weight (30.3%), and seen at an urban clinic setting (42.8%) versus a suburban (33.3%), rural (15.9%), or unknown clinic setting (13.9%). There was no significant difference in application of the diagnosis code by sex.

Of the children (n=15,423) who had BP ≥95^th^ percentile at ≥1 visit, only 6,427 (41.6%) had a corresponding ICD-10 code (R03.0). Similar to above, children with BP ≥95^th^ percentile who had the diagnosis code, versus those who did not, were more likely to be older (80.8% of children ≥12 years vs. 26.0% of children ≤6 years), Black (45.9%) versus white race (42.5%) and white versus Hispanic/Latino race (34.4%), Hispanic/Latino (46.4%) versus white ethnicity (36.6%), classified with obesity (50.8%) versus normal weight (33.0%), and seen at an urban (47.4%) versus a suburban (38.0%), rural (18.0%), or unknown clinic setting (15.5%). There was no significant difference in application of the diagnosis code by sex.

Out of 11,675 children who attended at least 3 visits during the study period, 2,542 (22%) had BP ≥95^th^ percentile at ≥3 visits and thus met criteria for pHTN. Of those children, only 146 (5.7%) had a corresponding ICD-10 code (I10). Children who met criteria had greater odds of having a pHTN diagnosis code if they were older (11.4% of children ≥12 years vs. 1.4% of children ≤6 years), Unknown (3.5%) versus white race (5.1%), Unknown (3.5%) versus white ethnicity (6.1%), classified with obesity (7.3%) versus normal weight (3.2%), and seen at a suburban (5.1%), rural (6.4%), or unknown (10.0%) versus an urban clinic setting (5.5%). There were no significant differences in application of the diagnosis code by sex or other specified racial or ethnic categories.

### CDS tool use

Of the full sample (N=23,334), the CDS tool was used to calculate systolic BP percentile for 10,524 (45.1%) children. Predictors of CDS tool use are presented in Supplemental Material (Table S1). When the CDS tool was used to calculate BP percentile, the odds of a children having an ICD-10 code of R03.0 was significantly greater for both BP ≥90^th^ percentile (56.1% vs. 27.4%; OR: 3.39, 95%CI: 3.21, 3.59) and BP ≥95^th^ percentile (56.9% vs. 31.1%; OR: 2.91,

95%CI: 2.72, 3.11). Likewise, the odds of children with BP ≥95^th^ percentile at ≥3 visits having an ICD-10 code of I10 were significantly greater if the CDS tool was used (10.8% vs. 1.9%; OR: 6.18, 95%CI: 4.06, 9.40).

### Guideline-adherent management

Predictors of guideline-adherent management were similar between BP ≥90^th^ and ≥95^th^ percentile; thus, logistic regression results for children with BP ≥95^th^ percentile are presented in Table 3 and below. Results for BP ≥90^th^ percentile are presented in Supplemental Table S2.

**Table 3.** Predictors of receiving guideline-adherent management among children with elevated blood pressure readings (≥95th percentile)

| Variable | Anti-hypertensive medication |  |  | Lifestyle counseling |  |  | Referral |  |  |
| --- | --- | --- | --- | --- | --- | --- | --- | --- | --- |
|  | OR | 95% CI |  | OR | 95% CI |  | OR | 95% CI |  |
| age, years | **1.113** | **1.096** | **1.130** | **0.945** | **0.928** | **0.963** | **0.965** | **0.950** | **0.981** |
| sex (ref: male) | **0.541** | **0.464** | **0.631** | **1.867** | **1.557** | **2.237** | 1.083 | 0.940 | 1.248 |
| Race (ref: Non-Hispanic white) |  |  |  |  |  |  |  |  |  |
| Black | **0.667** | **0.541** | **0.822** | **1.313** | **1.026** | **1.681** | 0.795 | 0.622 | 1.017 |
| Hispanic/Latino | **0.601** | **0.368** | **0.980** | 1.514 | 0.902 | 2.542 | 0.757 | 0.554 | 1.036 |
| Asian | **0.224** | **0.116** | **0.431** | **6.417** | **2.572** | **16.011** | 0.698 | 0.416 | 1.172 |
| More than one race | 1.222 | 0.729 | 2.051 | 1.050 | 0.539 | 2.044 | 1.220 | 0.597 | 2.491 |
| Other | **0.360** | **0.139** | **0.934** | 2.064 | 0.751 | 5.671 | 0.564 | 0.134 | 2.373 |
| Unknown | 0.765 | 0.523 | 1.120 | 1.151 | 0.749 | 1.770 | **0.443** | **0.255** | **0.770** |
| Ethnicity (ref: Not Hispanic/Latino) |  |  |  |  |  |  |  |  |  |
| Hispanic/Latino | **0.308** | **0.250** | **0.379** | **2.385** | **1.868** | **3.045** | **1.290** | **1.027** | **1.620** |
| Unknown | 0.784 | 0.506 | 1.214 | 0.906 | 0.572 | 1.435 | 0.881 | 0.500 | 1.550 |
| Weight status (normal BMI) |  |  |  |  |  |  |  |  |  |
| Underweight | 0.836 | 0.468 | 1.492 | 0.708 | 0.436 | 1.151 | 0.733 | 0.464 | 1.157 |
| Overweight | 0.816 | 0.652 | 1.022 | **1.444** | **1.119** | **1.865** | **0.603** | **0.494** | **0.736** |
| Obese | 0.902 | 0.766 | 1.061 | **1.733** | **1.428** | **2.104** | **0.276** | **0.229** | **0.332** |
| Clinic setting (ref: Urban) |  |  |  |  |  |  |  |  |  |
| Suburban | 1.074 | 0.862 | 1.339 | 1.090 | 0.847 | 1.404 | **0.690** | **0.561** | **0.849** |
| Rural | **1.961** | **1.594** | **2.413** | 1.172 | 0.883 | 1.555 | **0.014** | **0.004** | **0.058** |
| Unknown | 0.583 | 0.264 | 1.283 | 1.106 | 0.512 | 2.386 | **0.036** | **0.005** | **0.269** |
| Constant | **0.088** | **0.064** | **0.121** | **8.040** | **5.549** | **11.649** | **0.136** | **0.098** | **0.190** |

**Bold** indicates a significant predictor in a logistic regression predicting that eligible children had received guideline-adherent management documented in the EHR.
Abbreviations: BP: blood pressure. EHR: electronic health record. pHTN: pediatric hypertension. OR: odds ratio. CI: confidence interval. BMI: body mass index.

**Table 4.** Predictors of attending a guideline-adherent follow-up visit among children with elevated blood pressure readings

| Variable | Attended a follow-up visit within 6 months<br>after BP reading $\geq 90$th and $< 95$th percentile <sup>a</sup> | | | Attended a follow-up visit within 1 month after BP<br>reading $\geq 95$th percentile <sup>b</sup> | | |
| --- | --- | --- | --- | --- | --- | --- |
|  | OR | 95% CI |  | OR | 95% CI |  |
| age, years | **1.069** | **1.054** | **1.085** | **1.099** | **1.089** | **1.110** |
| sex (ref: male) | 1.055 | 0.928 | 1.198 | **0.895** | **0.819** | **0.977** |
| Race (ref: Non-Hispanic white) |  |  |  |  |  |  |
| Asian | 0.644 | 0.395 | 1.050 | 0.973 | 0.722 | 1.310 |
| Black | 0.857 | 0.694 | 1.058 | **0.795** | **0.687** | **0.919** |
| Hispanic/Latino | **0.579** | **0.438** | **0.766** | **0.664** | **0.542** | **0.815** |
| More than one race | 1.166 | 0.589 | 2.309 | 0.813 | 0.500 | 1.321 |
| Other | 0.862 | 0.343 | 2.170 | 0.559 | 0.286 | 1.092 |
| Unknown | 1.058 | 0.693 | 1.614 | 0.842 | 0.644 | 1.099 |
| Ethnicity (ref: Not Hispanic/Latino) |  |  |  |  |  |  |
| Hispanic/Latino | 0.890 | 0.731 | 1.084 | 1.013 | 0.888 | 1.156 |
| Unknown | 0.708 | 0.475 | 1.055 | 0.983 | 0.733 | 1.317 |
| Weight status (normal BMI) |  |  |  |  |  |  |
| Underweight | **2.255** | **1.157** | **4.397** | 0.969 | 0.686 | 1.370 |
| Overweight | 0.963 | 0.797 | 1.164 | 0.936 | 0.817 | 1.072 |
| Obese | **1.339** | **1.161** | **1.544** | 1.082 | 0.979 | 1.195 |
| Clinic setting (ref: Urban) |  |  |  |  |  |  |
| Suburban | **0.741** | **0.618** | **0.889** | 0.917 | 0.810 | 1.040 |
| Rural | **0.755** | **0.590** | **0.968** | 0.914 | 0.783 | 1.067 |
| Unknown |  |  |  |  |  |  |
| Constant | **0.515** | **0.372** | **0.713** | **0.109** | **0.088** | **0.134** |

**Bold** indicates a significant predictor in a logistic regression predicting that eligible children attended a guideline-adherent follow-up visit.
<sup>a</sup> 2,269 out of 4,197 (54.1%) eligible children attended a follow-up visit.
<sup>b</sup> 2,598 out of 15,164 (17.1%) eligible children attended a follow-up visit.
Abbreviations: BP: blood pressure. EHR: electronic health record. pHTN: pediatric hypertension. OR: odds ratio. CI: confidence interval. BMI: body mass index

Antihypertensive medication was prescribed to 795 children (5.2%) with BP ≥95^th^ percentile. Children who were prescribed antihypertensive medication were more likely to be older, male, white race versus Black, Hispanic/Latino, Asian, and Other, non-Hispanic/Latino versus Hispanic/Latino ethnicity, and attending a clinic in a rural rather than urban setting. There was no significant difference in antihypertensive medication prescriptions by weight status.

Lifestyle counseling rates were high among children with BP ≥95^th^ percentile (n=14,841, 96.2%). Children who received lifestyle counseling were more likely to be younger (98.6% of children ≤6 years vs. 96.8% of children ≥12 years), female (97.4% vs. male 95.3%), Asian (99.0%) or Black (95.1%) versus white race (96.4%), Hispanic/Latino (97.5%) versus white ethnicity (95.0%), and classified with overweight (96.7%) or obesity (97.0%) versus normal weight (95.4%). There was no significant difference in lifestyle counseling by clinic setting.

Rates of referrals for specialty services related to elevated BP were (n=848, 5.5%) among children with BP ≥95^th^ percentile. Children who received a referral were more likely to be younger (7.9%% of children ≤6 years vs. 4.5% of children ≥12 years), white (6.1%) versus Unknown race (3.9%), Hispanic/Latino (6.8%) versus non-Hispanic/Latino ethnicity (4.2%), classified with normal weight (8.9%) versus overweight (5.5%) or obesity (2.5%), and attend a clinic in an urban (6.7%) versus a suburban (5.0%), rural (0.1%), or unknown setting (0.3%). There was no significant difference in referrals by sex or other specified racial categories.

### Guideline-adherent follow-up

A total of 4,197 children with BP 90-94^th^ percentile were eligible for the follow-up visit analyses. Of those children, 2,269 (54.1%) attended a follow-up visit within 6 months. Children who attended a guideline-adherent follow-up visit were more likely to be older (58.8% of children ≥12 years vs. 40.9% of children ≤6 years), white (55.0%) versus Hispanic/Latino race (44.0%), be classified as underweight (65.9%) or obese (58.3%) versus normal weight (49.1%), and attend a clinic in an urban (55.6%) versus a suburban (48.6%) or rural setting (49.9%). There were no significant differences in 6-month follow-up visit attendance by sex or ethnicity.

Of 15,164 children with BP ≥95^th^ percentile who were eligible for the follow-up visit analyses, 2,598 (17.1%) attended a follow-up visit within 1 month. Children who attended a guideline-adherent follow-up visit were more likely to be male (18.1%) than female (15.9%), older (26.7% of children ≥12 years vs. 10.6% of children ≤6 years), and white (18.4%) versus Black (15.7%) or Hispanic/Latino race (13.7%). There were no significant differences in 1-month follow-up visit attendance by ethnicity, weight status, or clinic setting.

## Discussion

With the publication of the 2017 CPG for the diagnosis and management of pHTN, the rate of children who meet diagnostic criteria for pHTN has increased.^5,7^ To our knowledge, this is the first analysis to examine the rates of guideline-adherent pHTN diagnosis and management documented in the EHR using the 2017 CPG. Compared to a prior study in this population using the Fourth Report,^12^ guideline-adherent diagnosis rates for pHTN using the 2017 CPG were considerably lower (42% vs. 6%) while the rates of children diagnosed with pre-hypertension/elevated BP without a diagnosis of hypertension were higher (17% vs. 38%). Consistent with prior studies, children who were male, older (≥13 years old), and classified with overweight or obesity were more likely to be diagnosed with pHTN or elevated BP.^1,9,24^ Moreover, children seen in urban-based clinics were more likely to have a guideline-adherent diagnosis code for elevated BP, but not pHTN.

Behavioral changes are the first-line intervention recommended for treating elevated BP among children.^4^ Accordingly, >95% of patients in this sample received some form of lifestyle counseling. Children classified with overweight or obesity had 33% or 56% greater odds, respectively, of receiving lifestyle counseling. Approximately 5% of children meeting criteria for elevated BP/pHTN were prescribed antihypertensive medication or received a referral for potential BP-related specialty care. Interestingly, antihypertensive medications were more likely to be prescribed to children attending a clinic in a suburban or rural setting, while referrals were more likely to be given to those in in an urban setting. A recent survey of pediatric clinicians in the AllianceChicago network found that the majority of clinicians (71%) agreed that selecting and prescribing antihypertensive medications was a barrier to pHTN management, and most (74%) preferred to refer children with pHTN to a specialist for treatment.^25^ These findings highlight the need for pediatric clinicians, particularly in rural settings, to develop comfort and familiarity with antihypertensive medications when specialty services may not be as readily available to manage pHTN per the 2017 CPG. Furthermore, reducing barriers to accessing specialty care (e.g., telehealth) is critical for BP management in children.

Per the 2017 CPG,^4^ children with BP ≥90^th^ and ≥95^th^ percentile should attend a follow-up visit within 2 weeks and 6 months, respectively, for a follow-up BP reading. In the present sample, 54% of children with BP ≥90^th^ percentile attended a 6-month follow-up visit but only 17% of children with BP ≥95^th^ percentile attended a follow-up visit within 1 month, similar to a prior study (21%).^22^ Lack of patient follow-up inherently limits the ability of clinicians to confirm three occurrences of BP ≥95^th^ percentile and confidently diagnosis and treat pHTN. Specific strategies targeting patient attendance, employing home or ambulatory blood pressure monitoring,^26^ and reducing barriers to accessing specialty care, might improve follow-up and increase rates of pHTN diagnosis and subsequent management.

The findings from this study underscore the gap between evidence and practice of guideline implementation.^10,11^ To address this gap, evidence-based tools and implementation strategies are needed. CDS tools are often implemented to support clinicians and improve clinical care, such as increasing adherence to clinical guidelines. Unfortunately, the pHTN CDS tool implemented in AllianceChicago clinics was used on fewer than half the patients with elevated BP readings (≥90^th^ percentile), and uptake was significantly lower in rural compared to urban clinics. These findings are consistent with prior studies in pHTN and more broadly, indicating that these tools are effective but have low adoption.^21,27^ Thus, more comprehensive and multilevel implementation strategies are needed to increase uptake of effective CDS tools to promote guideline-adherent pHTN diagnosis and management. For example, a stakeholder advisory panel comprising AllianceChicago pediatricians and academic partners identified 18 discrete implementation strategies needed to improve adherence to the 2017 CPG, including education and training, workflow changes, and leadership support.^19^

### Limitations

First, the focus on high-risk children with at least one BP reading ≥90^th^ percentile limits the generalizability of these findings. Second, it is impossible to know whether the management options (medications, counseling, and referrals) were specific to elevated BP; for example, lifestyle counseling could have solely focused on weight management. Finally, the true prevalence of pHTN is unknown, as the diagnosis requires three instances of elevated BP (≥95^th^ percentile) and the rates of follow-up visit attendance were low.

## Conclusion

Elevated BP and pHTN continue to be underdiagnosed and undertreated among children seen in safety-net clinics. Use of a CDS tool to facilitate the recognition of elevated BP was underutilized. Appropriate diagnosis and management of pHTN per the 2017 CPG is critical for reducing pHTN-related morbidity and mortality among children, adolescents, and adults.^8,13,14,16,28^ More comprehensive implementation strategies are needed to address this research to practice gap, including strategies to support the implementation of health information technology tools.

## Data Availability

All data produced in the present study are available upon reasonable request to the authors.

## Notes

### Competing Interest Statement

The authors have declared no competing interest.

### Funding Statement

This study was funded by the National Heart, Lung, and Blood Institute, Grant/Award Number: R56HL148192; and the National Institute on Drug Abuse, Grant/Award Number: P30DA027828

### Author Declarations

IRB of Northwestern University Feinberg School fo Medicine gave ethical approval for this work.

## References

1. Bell CS, Samuel JP and Samuels JA. Prevalence of Hypertension in Children. Hypertension. 2019;73:148–152.

2. Song P, Zhang Y, Yu J, Zha M, Zhu Y, Rahimi K and Rudan I. Global Prevalence of Hypertension in Children: A Systematic Review and Meta-analysis. JAMA Pediatrics. 2019;173:1154–1163.

3. Skinner AC, Ravanbakht SN, Skelton JA, Perrin EM and Armstrong SC. Prevalence of Obesity and Severe Obesity in US Children, 1999-2016. Pediatrics. 2018;141.

4. Flynn JT, Kaelber DC, Baker-Smith CM, Blowey D, Carroll AE, Daniels SR, de Ferranti SD, Dionne JM, Falkner B, Flinn SK, Gidding SS, Goodwin C, Leu MG, Powers ME, Rea C, Samuels J, Simasek M, Thaker VV and Urbina EM. Clinical Practice Guideline for Screening and Management of High Blood Pressure in Children and Adolescents. Pediatrics. 2017;140:e20171904.

5. Yang L, Kelishadi R, Hong YM, Khadilkar A, Nawarycz T, Krzywińska-Wiewiorowska M, Aounallah-Skhiri H, Motlagh ME, Kim HS, Khadilkar V, Krzyżaniak A, Romdhane HB, Heshmat R, Chiplonkar S, Stawinska-Witoszynska B, Ati JE, Qorbani M, Kajale N, Traissac P, Ostrowska-Nawarycz L, Ardalan G, Ekbote V, Zhao M, Heiland EG, Liang Y and Xi B. Impact of the 2017 American Academy of Pediatrics Guideline on Hypertension Prevalence Compared With the Fourth Report in an International Cohort. Hypertension. 2019;74:1343–1348.

6. The fourth report on the diagnosis, evaluation, and treatment of high blood pressure in children and adolescents. Pediatrics. 2004;114:555–76.

7. Al Kibria GM, Swasey K, Sharmeen A and Day B. Estimated Change in Prevalence and Trends of Childhood Blood Pressure Levels in the United States After Application of the 2017 AAP Guideline. Prev Chronic Dis. 2019;16:E12.

8. Khoury M, Khoury P, Bazzano L, Burns TL, Daniels S, Dwyer T, Ikonen J, Jacobs DR, Jr., Juonala M, Kähönen M, Prineas R, Raitakari OT, Steinberger J, Venn A, Viikari J, Woo JG, Sinaiko A and Urbina EM. Prevalence Implications of the 2017 American Academy of Pediatrics Hypertension Guideline and Associations with Adult Hypertension. J Pediatr. 2021.

9. Hansen ML, Gunn PW and Kaelber DC. Underdiagnosis of Hypertension in Children and Adolescents. JAMA. 2007;298:874–879.

10. Patel ND, Newburn A, Brier ME and Chand DH. Pediatric Hypertension: Are Pediatricians Following Guidelines? J Clin Hypertens (Greenwich). 2016;18:1230–1234.

11. Rao G, Naureckas S, Datta A, Mohanty N, Bauer V, Padilla R, Rittner SS, Tilmon S and Epner P. Pediatric hypertension: diagnostic patterns derived from electronic health records. Diagnosis (Berl). 2018;5:157–160.

12. Moin A, Mohanty N, Tedla YG, Carroll AJ, Padilla R, Langman CB and Smith JD. Under-recognition of pediatric hypertension diagnosis: Examination of 1 year of visits to community health centers. J Clin Hypertens (Greenwich). 2021;23:257–264.

13. Khoury M and Urbina EM. Cardiac and Vascular Target Organ Damage in Pediatric Hypertension. Front Pediatr. 2018;6:148.

14. Urbina EM, Mendizábal B, Becker RC, Daniels SR, Falkner BE, Hamdani G, Hanevold C, Hooper SR, Ingelfinger JR, Lanade M, Martin LJ, Meyers K, Mitsnefes M, Rosner B, Samuels J and Flynn JT. Association of Blood Pressure Level With Left Ventricular Mass in Adolescents. Hypertension. 2019;74:590–596.

15. Dawson AE, Kallash M, Spencer JD and Wilson CS. The pressure’s on: understanding neurocognitive and psychological associations with pediatric hypertension to inform comprehensive care. Pediatr Nephrol. 2021.

16. Du T, Fernandez C, Barshop R, Chen W, Urbina EM and Bazzano LA. 2017 Pediatric Hypertension Guidelines Improve Prediction of Adult Cardiovascular Outcomes. Hypertension. 2019;73:1217–1223.

17. Theodore RF, Broadbent J, Nagin D, Ambler A, Hogan S, Ramrakha S, Cutfield W, Williams MJ, Harrington H, Moffitt TE, Caspi A, Milne B and Poulton R. Childhood to Early-Midlife Systolic Blood Pressure Trajectories: Early-Life Predictors, Effect Modifiers, and Adult Cardiovascular Outcomes. Hypertension. 2015;66:1108–15.

18. Bello JK, Mohanty N, Bauer V, Rittner SS and Rao G. Pediatric Hypertension: Provider Perspectives. Global pediatric health. 2017;4:2333794X17712637–2333794X17712637.

19. Knapp AA, Carroll AJ, Mohanty N, Fu E, Powell BJ, Hamilton A, Burton ND, Coldren E, Hossain T, Limaye DP, Mendoza D, Sethi M, Padilla R, Price HE, Villamar JA, Jordan N, Langman CB and Smith JD. A stakeholder-driven method for selecting implementation strategies: a case example of pediatric hypertension clinical practice guideline implementation. Implement Sci Commun. 2022;3:25.

20. Meisner JK, Yu S, Lowery R, Liang W, Schumacher KR and Burrows HL. Clinical Decision Support Tool for Elevated Pediatric Blood Pressures. Clinical Pediatrics. 2022;61:428–439.

21. Vuppala S and Turer CB. Clinical Decision Support for the Diagnosis and Management of Adult and Pediatric Hypertension. Curr Hypertens Rep. 2020;22:67.

22. Daley MF, Sinaiko AR, Reifler LM, Tavel HM, Glanz JM, Margolis KL, Parker E, Trower NK, Chandra M, Sherwood NE, Adams K, Kharbanda EO, Greenspan LC, Lo JC, O’Connor PJ and Magid DJ. Patterns of care and persistence after incident elevated blood pressure. Pediatrics. 2013;132:e349–55.

23. Stata Statistical Software: Release 17 [computer program]. College Station, TX: StataCorp LLC; 2021.

24. Rerksuppaphol L and Rerksuppaphol S. Prevalence and Risk Factors of Hypertension in Schoolchildren from Central Thailand: A Cross-Sectional Study. Int J Prev Med. 2021;12:28.

25. Carroll AJ, Tedla YG, Mohanty N, Padilla R, Moin A, Langman CB and Smith JD. Correlates of Adherence to the 2017 Clinical Practice Guidelines for Pediatric Hypertension in Safety-Net Clinics: A Two-Year Cross-Sectional Study. submitted for publication.

26. Peterson CG and Miyashita Y. The Use of Ambulatory Blood Pressure Monitoring As Standard of Care in Pediatrics. Frontiers in Pediatrics. 2017;5.

27. Kouri A, Yamada J, Lam Shin Cheung J, Van de Velde S and Gupta S. Do providers use computerized clinical decision support systems? A systematic review and meta-regression of clinical decision support uptake. Implementation Science. 2022;17:21.

28. Urbina EM, Khoury PR, McCoy C, Daniels SR, Kimball TR and Dolan LM. Cardiac and vascular consequences of pre-hypertension in youth. J Clin Hypertens (Greenwich). 2011;13:332–42.

