## Supplemental Material for "Correlates of Adherence to the 2017 Clinical Practice Guidelines for Pediatric Hypertension in Safety-Net Clinics: A Two-Year Cross-Sectional Study"

For

**Methods**

**Guideline-adherent management**

Antihypertensive medications extracted from the AllianceChicago EHR included Ace Inhibitors (Lisinopril, Enalapril, Captopril), Ace receptor blockers (Losartan, Valsartan), Calcium channel blockers (Amlodipine, Isradipine, Nifedipine), Beta blockers (Propranolol, Metoprolol, Labetalol, Carvedilol, Atenolol), Alpha Blockers (Doxazosin), Alpha 2 agonist (Clonidine), Diuretics, (Furosemide, Chlorothiazide, Metolazone, Spironolactone, Amiloride), and Vasodilator.

**Results**

**CDS tool use**

Of the full sample (N=23,334), the CDS tool was used to calculate the systolic BP percentile for 10,524 (45.1%) children. The CDS tool was more likely to be used for children who were female (OR: 1.139, 95%CI: 1.075, 1.205), younger (OR: 0.809, 95%CI: 0.803, 0.815), Hispanic/Latino (OR: 1.204, 95%CI: 1.103, 1.314) or unknown ethnicity (OR: 1.222, 95%CI: 1.021, 1.462), and seen in an urban setting compared to those seen in a rural (OR: 0.603, 95%CI: 0.544, 0.668) or unknown clinic setting (OR: 0.542, 95%CI: 0.429, 0.685). There was no significant difference in the use of the CDS tool by weight status.

**Supplemental Table S1**. Predictors of having clinical decision support (CDS) tool-calculated blood pressure percentile

|  | Systolic blood pressure | | | Diastolic blood pressure | | |
| --- | --- | --- | --- | --- | --- | --- |
| Variable | OR | 95% CI | | OR | 95% CI | |
| age, years | **0.809** | **0.803** | **0.815** | **0.809** | **0.803** | **0.815** |
| sex (ref: male) | **1.139** | **1.075** | **1.205** | **1.139** | **1.075** | **1.205** |
| Race (ref: Non-Hispanic white) | |  |  |  |  |  |
| Asian | 1.175 | 0.981 | 1.408 | 1.174 | 0.980 | 1.406 |
| Black | 1.060 | 0.964 | 1.165 | 1.060 | 0.964 | 1.166 |
| Hispanic/Latino | **0.595** | **0.525** | **0.674** | **0.595** | **0.525** | **0.674** |
| More than one race | 0.783 | 0.607 | 1.011 | 0.782 | 0.606 | 1.009 |
| Other | 1.323 | 1.000 | 1.750 | 1.322 | 0.999 | 1.749 |
| Unknown/Not specified | 1.135 | 0.957 | 1.346 | 1.134 | 0.956 | 1.345 |
| Ethnicity (ref: Not Hispanic/Latino) | |  |  |  |  |  |
| Hispanic/Latino | **1.204** | **1.103** | **1.314** | **1.203** | **1.102** | **1.313** |
| Unknown/Not specified | **1.222** | **1.021** | **1.462** | **1.221** | **1.020** | **1.461** |
| Weight status (normal BMI) | |  |  |  |  |  |
| Underweight | 1.046 | 0.859 | 1.275 | 1.045 | 0.857 | 1.273 |
| Overweight | 0.943 | 0.868 | 1.025 | 0.943 | 0.868 | 1.025 |
| Obese | 1.010 | 0.946 | 1.078 | 1.009 | 0.945 | 1.077 |
| Clinic setting (ref: Urban) |  |  |  |  |  |  |
| Suburban | 1.081 | 0.995 | 1.173 | 1.080 | 0.995 | 1.172 |
| Rural | **0.603** | **0.544** | **0.668** | **0.603** | **0.545** | **0.668** |
| Unknown/Not specified | **0.542** | **0.429** | **0.685** | **0.542** | **0.429** | **0.685** |
| Constant | **4.176** | **3.754** | **4.646** | **4.186** | **3.762** | **4.657** |

**Bold** indicates a significant predictor in a logistic regression predicting that a clinician used the CDS tool to calculate blood pressure percentile in the EHR.

Abbreviations: OR: odds ratio. CI: confidence interval. BMI: body mass index.

**Supplemental Table S2**. Predictors of receiving guideline-adherent intervention among children with BP ≥90th percentile

|  | Anti-hypertensive medication | | | Lifestyle counseling | | | Referral | | |
| --- | --- | --- | --- | --- | --- | --- | --- | --- | --- |
| Variable | OR | 95% CI | | OR | 95% CI | | OR | 95% CI | |
| age, years | **1.117** | **1.101** | **1.134** | **0.942** | **0.927** | **0.958** | **0.964** | **0.950** | **0.977** |
| sex (ref: male) | **0.571** | **0.496** | **0.657** | **1.753** | **1.501** | **2.046** | 1.048 | 0.935 | 1.176 |
| Race (ref: Non-Hispanic white) | | |  |  |  |  |  |  |  |
| Black | **0.198** | **0.104** | **0.378** | **5.077** | **2.535** | **10.166** | 0.862 | 0.577 | 1.288 |
| Hispanic/Latino | **0.707** | **0.584** | **0.856** | **1.280** | **1.032** | **1.587** | 0.835 | 0.682 | 1.022 |
| Asian | **0.624** | **0.401** | **0.970** | 1.198 | 0.806 | 1.782 | 0.720 | 0.560 | 0.927 |
| More than one race | 1.183 | 0.732 | 1.913 | 1.186 | 0.656 | 2.144 | 1.257 | 0.711 | 2.221 |
| Other | **0.351** | **0.149** | **0.831** | **3.655** | **1.384** | **9.656** | 0.511 | 0.159 | 1.641 |
| Unknown | 0.845 | 0.593 | 1.204 | 1.117 | 0.761 | 1.639 | **0.456** | **0.288** | **0.723** |
| Ethnicity (ref: Not Hispanic/Latino) | | |  |  |  |  |  |  |  |
| Hispanic/Latino | **0.327** | **0.270** | **0.395** | **2.255** | **1.823** | **2.789** | **1.348** | **1.117** | **1.627** |
| Unknown | 0.720 | 0.482 | 1.076 | 1.021 | 0.680 | 1.531 | 0.853 | 0.543 | 1.339 |
| Weight status (normal BMI) | | |  |  |  |  |  |  |  |
| Underweight | 0.880 | 0.516 | 1.499 | **0.630** | **0.419** | **0.945** | 0.909 | 0.640 | 1.292 |
| Overweight | 0.909 | 0.742 | 1.112 | **1.327** | **1.068** | **1.648** | **0.663** | **0.567** | **0.776** |
| Obese | 0.998 | 0.860 | 1.159 | **1.561** | **1.319** | **1.848** | **0.277** | **0.237** | **0.324** |
| Clinic setting (ref: Urban) | |  |  |  |  |  |  |  |  |
| Suburban | 1.052 | 0.855 | 1.295 | 1.051 | 0.844 | 1.308 | **0.623** | **0.522** | **0.743** |
| Rural | **2.111** | **1.747** | **2.550** | 1.175 | 0.913 | 1.512 | **0.010** | **0.003** | **0.041** |
| Unknown | 0.808 | 0.401 | 1.629 | 0.569 | 0.320 | 1.011 | **0.030** | **0.004** | **0.216** |
| Constant | **0.054** | **0.040** | **0.073** | **11.398** | **8.241** | **15.765** | **0.131** | **0.100** | **0.171** |

**Bold** indicates a significant predictor in a logistic regression predicting that eligible children had received guideline-adherent intervention documented in the EHR.

Abbreviations: BP: blood pressure. EHR: electronic health record. pHTN: pediatric hypertension. OR: odds ratio. CI: confidence interval. BMI: body mass index.
